# Rethinking a hybrid malaria chemoprevention delivery strategy for children in sub-perennial settings: integrating age- and seasonally-targeted delivery

**DOI:** 10.1101/2025.04.03.25325165

**Authors:** Swapnoleena Sen, David Schellenberg, Melissa A Penny

## Abstract

**Background:** The World Health Organization recommends perennial malaria chemoprevention (PMC), generally using sulphadoxine-pyrimethamine (SP) to children at high risk of severe *P. falciparum* malaria. Currently, PMC is given up to age two in perennial transmission settings. However, no recommendation exists for perennial settings with seasonal variation in transmission intensity, recently categorized as ‘sub-perennial’. Tailored chemoprevention strategies are needed to protect children during seasons and ages of highest malaria risk. The seasonal dimension must adequately cover seasonally increased risk periods, alongside interventions that address year-round, lower intensity transmission. We propose a hybrid malaria chemoprevention (HMC) strategy, integrating two delivery components: 1) existing PMC, and 2) additional monthly SP doses during the higher-risk rainy season, ensuring a one-month gap between any two doses.

**Methods:** Using a validated individual-based malaria model combined with pharmacological models of drug action (OpenMalaria), we examined the potential public health impact of the proposed HMC (for children 03-24 months), and an age-expanded HMC (referred to as HMC+, for children 03-36 months), under different drug sensitivity, coverage, and prevalence (5-70%) assumptions.

**Results:** HMC and HMC+ demonstrated a median (interquartile range) of 2.1 (1.6–2.6), 2.9 (2.2-3.6) times higher efficacy (relative fold increase in burden averted) compared to only PMC against clinical, and 2.0 (0.6–3.4), 3.3 (0.8-5.8) against severe cases, respectively, in children under age three. This led to a median protective efficacy of 31.8% (25.4-38.2%), 44.9% (36.9-52.9%) against clinical, and 16.1% (7.0-25.2%), 26.4% (14.4-38.4%) against severe cases by HMC and HMC+ respectively, across the prevalence, drug sensitivity, and coverage assumptions. We found positive net impact for children under age five years, outweighing a limited potential of delayed malaria across settings.

**Conclusion:** Substantially increased public health benefits might be achieved by adding seasonally-targeted chemoprevention to current PMC in sub-perennial malaria transmission settings. Effectiveness-implementation studies should generate empirical evidence of public health impact including on the disease burden averted, safety, and cost-effectiveness of the hybrid approach. Such studies should also explore determinants of implementation success including operational feasibility, and acceptability of proposed dosing strategies which will facilitate deployment decisions.

## Background

Traditionally, the malaria community has thought in terms of ‘seasonal malaria’, with intense transmissions for three to five months per year and ‘perennial transmission’, which implies malaria is transmitted year-round fairly constantly. Currently, the World Health Organization (WHO)’s guidelines for malaria chemoprevention in children target these two endemic settings through contextual delivery strategies[<u>1</u>]. Perennial malaria chemoprevention (PMC) consists of administering repeated antimalarial treatment with sulphadoxine-pyrimethamine (SP) to young children (typically 03-24 months of age) in perennial *P. falciparum* malaria transmission settings. The age-targeted dosing time points are encouraged to align with local Expanded Program of Immunization (EPI) touchpoints. On the other hand, seasonal malaria chemoprevention (SMC) consists of three to five cycles of antimalarial treatment at least one month apart, usually to children under five years, administered during the peak transmission season in seasonal setting through fixed-point or door-to-door delivery[<u>1-3</u>].

However, many transmission settings do not fit into either the strictly seasonal or perfectly perennial. Most such settings have year-round transmission with seasonal variation in transmission intensity, leading to substantially higher transmission during certain months (such as, between December and April in parts of Mozambique). Recognizing this, WHO adopted a relatively new ‘sub-perennial’ malaria seasonality concept in 2021[<u>4</u>] However, to date, there are no chemoprevention recommendations specific to these areas[<u>1</u>] and particularly limited consideration to address how much seasonal variation warrants prioritising seasonally-targeted interventions. As such, SMC is usually not implemented in sub-perennial transmission settings, and PMC provides reduced protection if dosing does not coincide with the peak rainfall and parasite transmission timings[<u>5</u>].

We searched on PubMed (between 2000 and 2024, with search term “malaria” AND “sub-perennial” OR “subperennial”) but found no published study that explored any antimalarial strategy for such settings. However, through additional searches for perennial settings, we found results from one clinical study[<u>6</u>], a secondary analysis utilizing former intermittent preventive treatment in infants (IPTi) trial data[<u>7</u>], and one modelling[<u>8</u>] study that indicated promising potential benefits by administering seasonally-targeted monthly SP doses in children up to 24 months in settings now defined as sub-perennial. Up to three-fold greater efficacy was observed[<u>5</u>, <u>7</u>] for children who received their first two doses during the wet season compared to age-targeted dosing linked with the EPI schedule (75.2% vs 24.8% against clinical malaria). Similarly, higher efficacy per dose was predicted with monthly doses of SP in infants during the wet season only (26% at 24 months of age) compared with age-targeted IPTi-SP[<u>8</u>]. The efficacy further increased with seasonally targeted SP’s age-expansion up to 24 months (52% by 24 months of age). However, there are likely challenges to setting up and delivering seasonally-targeted SP[<u>1</u>], and currently, no such strategy is implemented.

Relying only on seasonally-targeted SP for the newly classified sub-perennial settings may hinder the adoption of either age-targeted SP (PMC) or the seasonally-targeted SP. In addition, research is needed to enhance PMC’s public health impact, including the benefit, cost, and coverage if any alternative regimen is needed to fit for different epidemiological or clinical contexts[<u>1</u>] Thus, we explored a new chemoprevention schedule to address these gaps via modelling. We propose that there is potential to reduce the childhood malaria burden by implementing a hybrid malaria chemoprevention (HMC) approach in sub-perennial settings by combining PMC administered throughout the year using the existing EPI platform and an additional four seasonally-targeted SP doses every year (likely through alternative delivery channels or by strengthening the capacity of EPI). We explored this by modelling the proposed HMC for children 03-24 months, consistent with the typical PMC age range[<u>1</u>] However, the risk of severe malaria remains high up to 36 months[<u>1</u>, <u>9</u>], particularly in settings with increasing seasonality[<u>9</u>] and additional EPI touch points occur in this age range (such as for booster doses of diphtheria and tetanus vaccines and the second dose of the measles vaccine)[<u>1</u>, <u>10</u>, <u>11</u>]. Thus, we also modelled an age-expanded HMC (referred here as HMC+, for children 03-36 months, including two more age-targeted dosing, and another rounds of seasonally-targeted dosing in the third year of life). Furthermore, it is crucial to monitor post-intervention effects when designing or expanding time-limited antimalarial intervention targeted for young children to estimate any potential interference with natural immunity acquisition[<u>12</u>] Therefore, we assessed any risk of delayed malaria and net program benefit for HMC and HMC+, in children up to five years of age.

Updating or designing new chemoprevention strategies needs careful planning, sustainable commitment, and communication of benefits to multiple stakeholders, including communities and families of the children. While this is primarily a modelling study, we additionally explored possible translation to public health practice in the light of implementation research[<u>13</u>, <u>14</u>]. While preliminary, we aimed to support more official community participation and any pilot implementation study design based on our understanding of the proposed intervention’s context[<u>13</u>] and to facilitate engagement of multidisciplinary research around a much-needed chemoprevention strategy. Altogether, this model-driven commentary aims to initiate dialogues based on quantitative evidence from representative scenarios and calls for generating empirical evidence on the safety, impact, feasibility, costs and acceptability of the proposed chemoprevention for protecting underserved children in sub-perennial settings.

## Methods

### Brief description of the model

We estimated the impact of chemoprevention strategies using our open-source, individual-based stochastic models of *P. falciparum* malaria epidemiology (OpenMalaria, https://github.com/SwissTPH/openmalaria/wiki)[<u>15</u>, <u>16</u>]. Different model variants describe varying assumptions of malaria pathophysiology, within-host parasite dynamics, the effects of comorbidity, heterogeneity of anti-malarial immunity acquisition and its decay. All models have been fitted to field data across sub-Saharan Africa, as described previously[<u>15</u>, <u>17</u>] (supplementary section 1.1, Table S1). We utilised a model variant embedding within-host parasite dynamics and drug PK/PD models[<u>17</u>] To model varying drug sensitivity to SP, we specified different thresholds for the half-maximal effective concentration (EC50): either full sensitivity against wild-type *P. falciparum*, or partial resistance against prevalent quadruple mutant genotypes (dhfr-51I, dhfr-59A, dhfr-108A, and dhps-437G in *Pfdhfr* and *Pfdhps* genes)[<u>18</u>, <u>19</u>].

### Simulation scenarios

We estimated the potential public health impacts, including any post-intervention effect of the proposed HMC compared to PMC alone. PMC doses align with age patterns of severe malaria as per current WHO recommendation (Figure 1a), and the seasonally-targeted dosing was informed by the clinical incidence pattern (Figure 1c). A one-month gap between any two doses was applied to avoid over-dosing[<u>1</u>]. Three SP dosing schedules were modelled under different drug sensitivity and chemoprevention coverage assumptions (Table 1): i) PMC (total seven EPI-linked SP doses spread between 03-24 months of age) ii) PMC+ (total nine EPI-linked SP doses spread between 03-36 months of age) iii) HMC (PMC, plus four monthly doses during the wet season for children 03-24) and iv) HMC+ (PMC+, plus four monthly doses during the wet season for children 03-36 months). During rainy season, seasonally-targeted SP doses take precedence and any age-targeted PMC doses are dropped if the schedules overlap.

**Figure 1.**
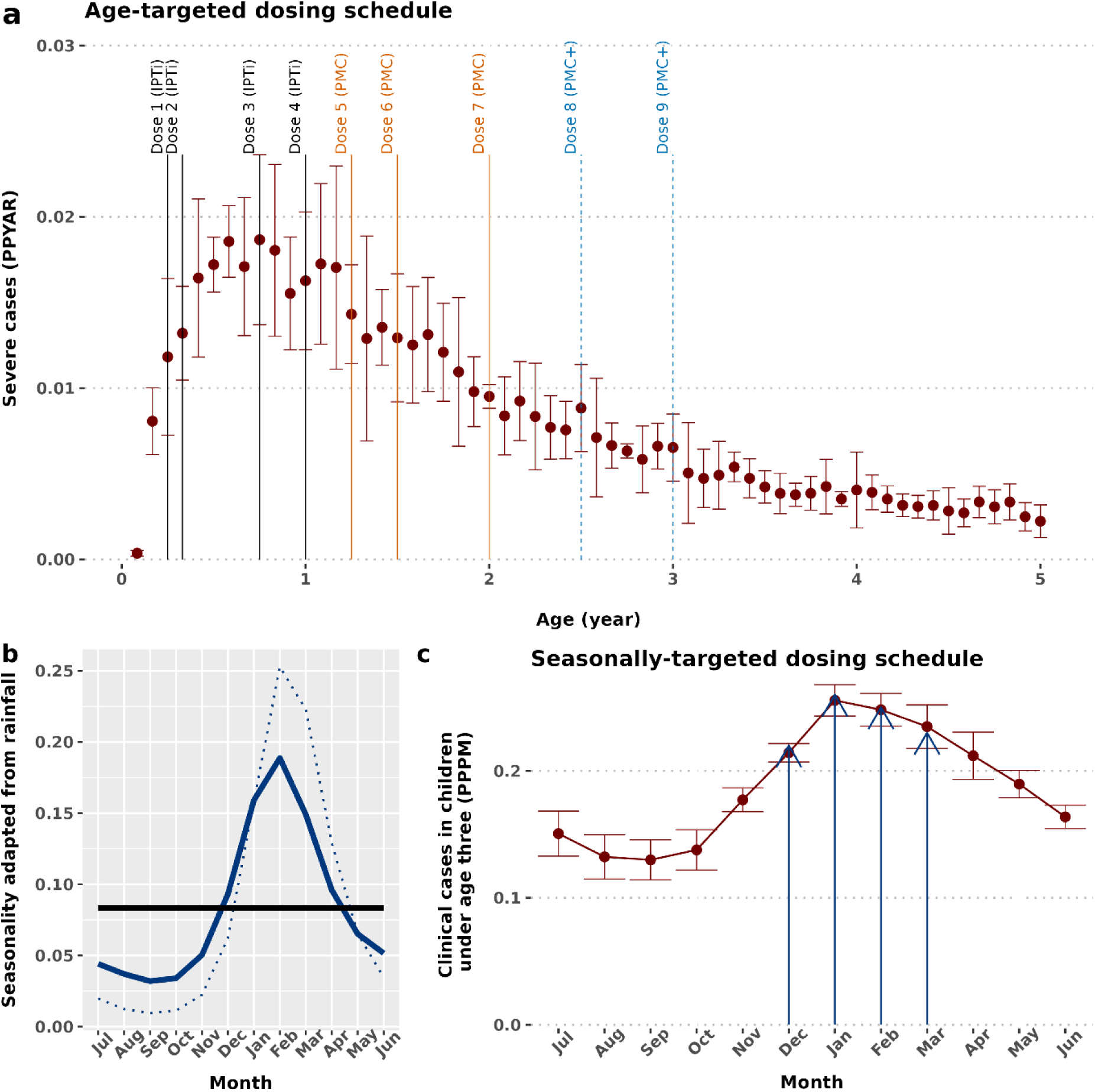
Proposed hybrid malaria chemoprevention with sulphadoxine-pyrimethamine (SP). **The top panel (a) depicts PMC and PMC+ dose schedule based on the age-pattern of severe malaria (median and interquartile range), and potential contacts with routine expanded program of immunization.** Four SP doses at 03, 04, 09 and 12 months of age (as per former IPTi, and also given for current PMC and for an age-expanded PMC+) is depicted by the solid vertical black line, three SP doses at 15, 18 and 24 months of age (as per current PMC) are shown by the solid orange lines, and two additional SP doses at 30 and 36 months of age (as per age-expanded PMC+) are depicted by the light blue dashed lines. **The bottom left panel (b) shows an example of a seasonality pattern (reflecting percentage of average yearly rainfall distributed across months).** The solid black line denotes perfect perennial transmission, dotted blue line shows seasonality pattern based on average monthly rainfall data from Mozambique, and the solid blue line indicates the modelled representative sub-perennial malaria transmission (adjusted from the Mozambique rainfall pattern). **The bottom right panel (c) shows the proposed additional four seasonally-targeted dosing cycles based on the pattern of clinical malaria incidence over the year.** The blue arrows show the seasonally-targeted dosing time-points. PPPM: Per person per month; PPYAR: Per person per year at risk

**Table 1:** Overview of the proposed hybrid malaria chemoprevention in children.

| Variable | Description | Value |
| --- | --- | --- |
| **Seasonality**<br>**(transmission pattern)** | Distribution of malaria incidence throughout the year, correlated with average rainfall and temperature. | 1. Representative sub-perennial [20]<br>*(adapted from rainfall pattern in Mozambique)*<br>2. Archetypal, constant perennial |
| **Transmission intensity** | Entomological inoculation rate (EIR), interpreted as the number of infectious mosquito bites received yearly by a human host | 1. 4, 8, 16, 32, 64, 128, 256, 512, 1024 |
| ***Plasmodium falciparum* prevalence (*PfPR*<sub>2-10</sub>)** | Test-positive patent malaria cases among 2-10 year olds using microscopy diagnostic | 2. *PfPR*<sub>2-10</sub> calculated as model output based on patent infection |
| **Chemoprevention dosing schedule** | Dosing timing for combining PMC or PMC+ with additional seasonally-targeted SP as per WHO recommendations: the current WHO guidelines allow flexibility in terms of selecting number of doses to best suit the context, and additionally recommends at least one month gap between consecutive doses[1]. | **HMC**<br>• **PMC:** Seven SP doses at 3, 4, 9, 12, 15, 18, 24 months of age<br>• **Seasonally-targeted SP:** four monthly doses in December to March (age 03-24 months) |
|  |  | **HMC+**<br>• **PMC+:** Nine SP doses at 3, 4, 9, 12, 15, 18, 24, 30, 36 months of age<br>• **Seasonally-targeted SP:** four monthly doses in December to March (age 03-36 months) |
| **Chemoprevention coverage (%)** | Proportion of eligible children receiving a chemoprevention (PMC or PMC+ or HMC or HMC+) in each dosing cycle:<br>1. Full coverage—modelled to estimate maximum achievable impact, and the maximum risk of post-intervention effects<br>2. Reduced coverage—modelled to simulate minimum WHO vaccination targets in all district-level populations [21, 22] | 1. 100% (full coverage setting)<br>2. 80% coverage for each cycle (reduced coverage setting). This means on average less than 10% receive all hybrid SP doses (i.e. only 8.59% children receive all HMC, and 5.50% receive all HMC+ doses). |
| **Case-management coverage (%)** | Access to effective treatment against clinical malaria incidence, expressed as probability in a 14-day period | 1. Low: 10 %<br>2. Medium: 30 %<br>3. High: 50 % |
|  |  | 4. Very high: 80 % |
| **Deterministic diagnostic** | Microscopy diagnostics detecting parasite density above specified threshold | 40 parasites per $\mu\text{L}$ of blood [23] |
| **PK parameters for SP<sup>2</sup> (one compartment model) [19, 24]** | 1. Volume of distribution (L/kg)<br>2. Absorption rate constant (per day)<br>3. Elimination rate constant (per day)<br>4. Negligible concentration (mg/L) | 1. 0.29<br>2. 12.5<br>3. 0.12<br>4. 0.001 |
| **PD parameters for SP<sup>2</sup> (Michaelis-Menton kinetic<sup>3</sup>)** | 1. Maximum killing rate (per day)<br>2. Slope of the effect curve<br>3. Drug concentration at which half the maximum killing rate is achieved (EC50; mg/L). Impacted by parasite genotype (mutations in key genes that drive drug-sensitivity and the duration of chemoprevention) | 1. 2.3<br>2. 2.1<br>3. 2.4 for partially SP-resistant having duration of protection 35 days (quadruple mutation in *Pf dhfr* gene encoding dihydrofolate reductase+ *Pf dhps* gene encoding dihydropteroate synthase) [7, 18, 25])<br>4. 0.5 for SP-sensitive having duration of protection 42 days [7, 18, 25] |

Pre-validated EC50 values were used to model SP’s prophylactic period under different drug sensitivity assumption[18, 19], reducing from 42 days in SP-sensitive setting to 35 days in partially SP-resistant setting[18]

The representative sub-perennial seasonality in our study approximates the transmission pattern in Mozambique[<u>20</u>]. The average monthly rainfall from Mozambique was adapted within OpenMalaria (https://swisstph.github.io/openmalaria/fourier) to demarcate from a strictly seasonal setting by ensuring less than 60% rainfall during a consecutive four month period. The distribution of entomological inoculation rate (EIR) was correlated to it in the model[<u>1-3</u>] (Figure 1b, supplementary section 1.2, and supplementary Table S2). However, since rainfall may vary between perennial to sub-perennial patterns over time, we also modelled archetypal perennial settings by a uniform year-round distribution of EIR (Figure 1b). The combination of setting characteristics are summarized in Table 1, and our simulation scenarios are a full factorial design of these characteristics with 10 stochastic realizations per scenario.

### Estimation of protective efficacy, and additional burden averted by the hybrid strategies

We reported results after five years from program rollout to capture both the impact during the intervention and for post-intervention follow-up ages[<u>12</u>]. The protective efficacy (at 100% coverage) was calculated as the relative reduction of incidence rate in the intervention group compared to the control group (absence of PMC). Similar was calculated for lower coverages. We also calculated the relative fold increase in burden averted by HMC or HMC+ compared to PMC as the effectiveness of HMC divided by the effectiveness of PMC.

### Estimation of post-intervention effects

Any post-intervention effects including: i) the age-pattern of clinical and severe cases per age, and ii) net impact (measured by the cumulative incidence by age) were estimated as per the WHO recommendation [<u>12</u>] up to age five. We report this result assuming full program coverage to predict the maximum extent of potential post-intervention effects.

### Statistical analysis

The relative contribution of different setting characteristics across the parameter ranges (indicated in table 1) on the protective effectiveness was assessed by applying multiple factorial ANOVA test using R Package CGPfunctions[<u>26</u>]. The contribution of individual setting characteristics after controlling for all factors was extracted from the partial eta squared values.

### Validation of SP model and impact estimates

Our hybrid malaria chemoprevention study follows a PMC modelling study that predicted the public health impact of a proposed age-expanded PMC schedule[<u>25</u>]. The likely parasite life-stage specific mode of action of SP and validation of our model estimated effect size (protective efficacy against clinical and severe malaria) against empirical data from a wide range of epidemiological and clinical settings across Africa[<u>20</u>, <u>27</u>] was described previously in detail[<u>25</u>].

### Exploration of implementation study designs and determinants of outcome

We acknowledge likely know-do gaps and the complexity of implementing any new chemoprevention recommendation[<u>28</u>]. Thus, we explored potential implementation designs to support pilot studies for assessing the utility and feasibility of our proposed HMC or HMC+ schedule. An initial discussion with implementation science researchers followed by a targeted literature search[<u>13</u>] was carried out to identify potential study designs that could facilitate a faster transition from proposal to practice. Additionally, the consolidated framework of implementation research (CFIR)[<u>14</u>] was considered as a reference to list likely determinants (such as the local and broader setting characteristics and stakeholders) of implementation outcomes (supplementary section 1.3 and Table S3). These factors were prospectively mapped to our proposed chemoprevention context based on the publicly available information of a recent PMC scale-up project (MULTIPLY)[<u>29</u>].

## Results

### Protective effectiveness

Consistent with earlier studies[<u>6-8</u>], our results indicated substantial public health benefits of seasonally-targeted SP doses (Figure 2a, 2b). We predicted a maximum median efficacy of 28.7%(25.1-32.3%) and 40.7% (36.2-45.2%) against clinical, and 14.4% (6.0-22.9%), and 18.1% (8.5%-27.8%) against severe malaria was achieved by HMC, and HMC+ respectively, in the first three of life in partially SP-resistant setting (Figure 2a). These values increased to 34.8% (31.5-38.1%), 48.4% (43.7-53.1%) against clinical, and 18.1% (8.5-27.8%), 29.4% (17.4-41.5%) against severe disease, under HMC and HMC+, respectively in SP-sensitive settings. Thereby, HMC and HMC+ increased the impact against clinical malaria by 2.1 (1.6 -2.6) times and 2.9 (2.2-3.6) times relative to PMC alone (relative fold increase in burden averted). Corresponding values were 2.0 (0.6-3.4) and 3.3 (0.8-5.8) against severe malaria (Figure 2b).

**Figure 2.**
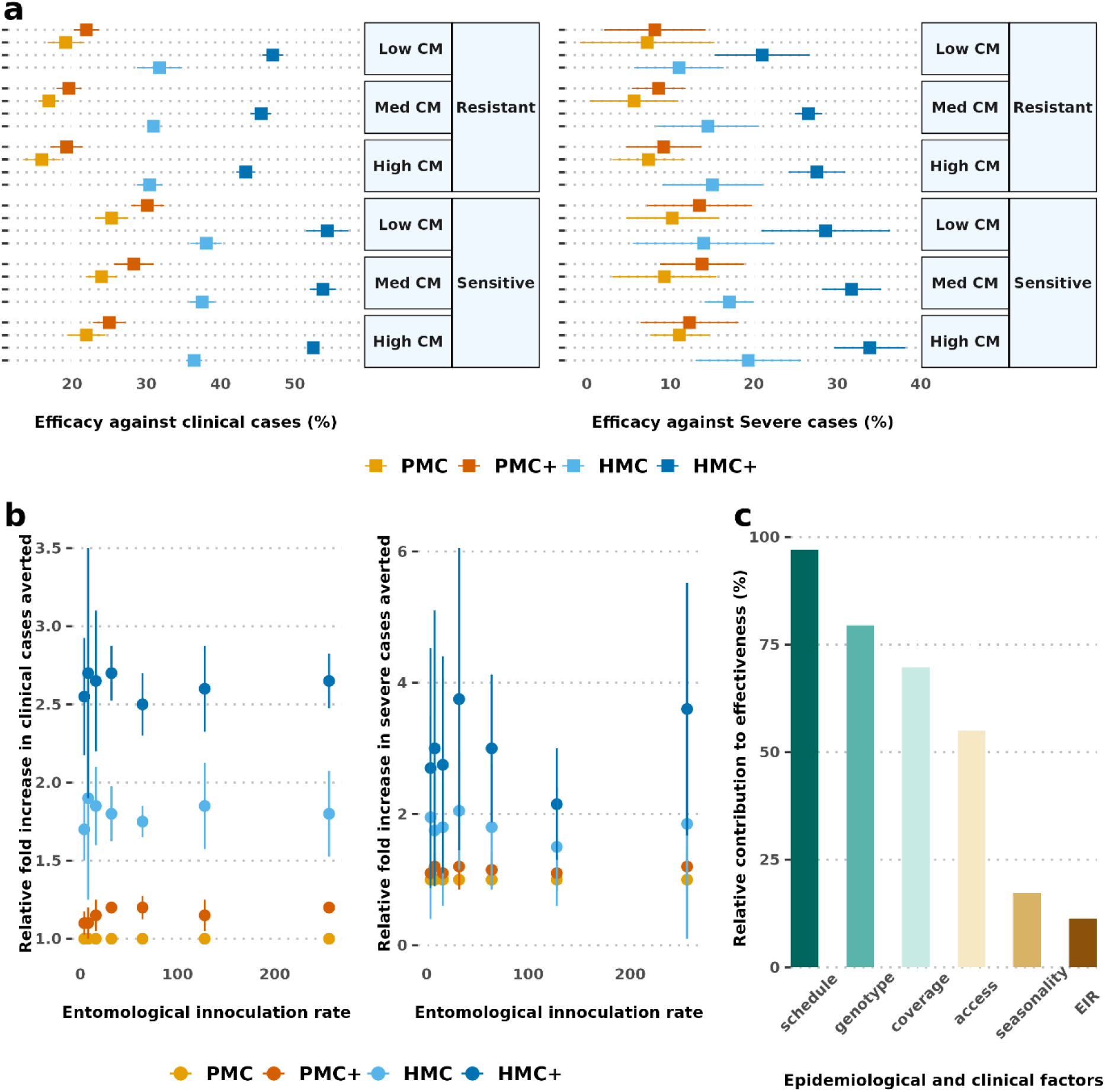
Protective efficacy and relative fold increase in burden averted by proposed hybrid dosing strategies in children under age three. **Panel a depicts the median (interquartile range) of protective efficacy across settings.** Results are shown for moderate to high transmission (baseline annual prevalence *Pf*PR_2-10_ 5-70%) and different healthcare strength setting**. Panel b depicts the relative fold increase in burden averted compared to PMC alone, while panel c shows the relative contribution of the setting and intervention characteristics on achievable effectiveness.** The relative fold increase in burden averted here is shown for full coverage as an exemplar of best-case scenario. CM: access to case management; EIR: entomological inoculation rate; HMC: hybrid malaria chemoprevention. HMC+: age-expanded HMC; PMC: perennial malaria chemoprevention; PMC+: age-expanded perennial malaria chemoprevention.

The statistical analysis showed that the dosing schedule will likely drive this increased protective effectiveness across epidemiological and clinical settings (Figure 2c). Consistent with earlier studies[<u>28</u>, <u>30</u>], our results demonstrated that the protection remained largely sustained in partially SP-resistant settings (such as quadruple dhfr-51I, dhfr-59A, dhfr-108A, and dhps-437G mutations in *Pf*dhfr and *Pf*dhps genes that reduced the prophylactic period to 35 days from 42 days in the sensitive setting)[<u>18</u>].

We also predicted that the enhanced impact (relative fold increase in burden averted) of hybrid chemoprevention remains high in reduced coverage assumptions (Supplementary Figure S1). These values reached 1.7 (1.2-2.2), and 2.5 (1.9-3.1) against clinical, and 1.7 (0.5-2.9) and 2.7 (0.8-4.6) against severe cases by HMC and HMC+ with 80% coverage in each dosing cycles (leading to 8.59% children to receive all HMC, and 5.50% children to receive all HMC+ doses). The number of doses contributed most to increasing effectiveness, followed by coverage against clinical malaria. Coverage was more important than age-expansion against severe cases for PMC, but not for HMC.

### Overall net intervention impact

The net impact (i.e. total intervention and post-intervention effects)[<u>12</u>] is predicted to be positive when children up to age five were followed up. This indicates limited potential of delayed malaria for HMC and HMC+ (Figure. 3a, 3b). Total impact increased in higher prevalence settings, especially when higher access to treatment was available. The net impact was also sustainable under reduced chemoprevention coverage assumption (Figure 3c, 3d). These results confirmed that the positive net impact of proposed schedules outweighs the limited potential of delayed malaria (supplementary Figure S2).

**Figure 3.**
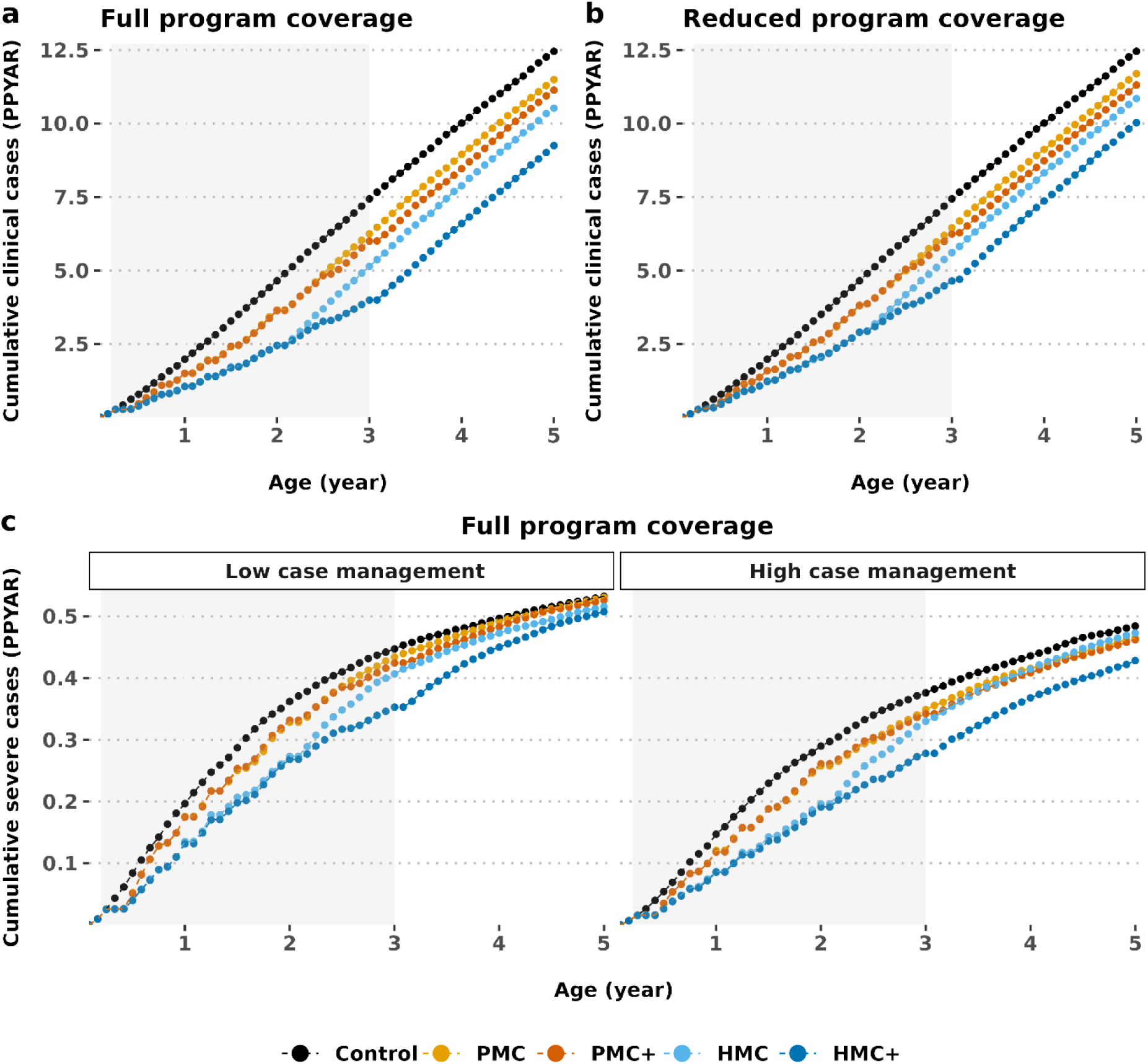
The net program impact expressed as cumulative malaria incidence in intervention and post-intervention ages. **The top panel (a and b) shows the net impact under full (100% in each dosing cycle of both age- and seasonally-targeted SP) vs. reduced program coverage assumptions (80% in each dosing cycle).** Results are shown in settings with *Pf*PR_2-10_ 30–39% (entomological inoculation rate 32 and 30% probability of access to case management in a 14 day post-diagnostic period). **The bottom panel (c) indicates the net impact in low (10% probability in a 14 day post-diagnostic period) vs. high access to treatment (50% probability in a 14 day post-diagnostic period) settings**, **under full chemoprevention coverage assumption**. HMC: hybrid malaria chemoprevention. HMC+: age-expanded HMC; PMC: perennial malaria chemoprevention; PMC+: age-expanded perennial malaria chemoprevention; PPYAR: Per person per year at risk.

### Reflection on implementation designs

Prospective pilot studies employing hybrid study designs that simultaneously focus on assessing clinical effectiveness and implementation are likely most useful and suitable for generating empirical data on the proposed HMC and HMC+ strategy[<u>13</u>]. As such, a hybrid type 2 effectiveness-implementation study that includes both clinical and implementation outcomes will likely generate quantitative evidence of effectiveness, cost-effectiveness, safety and any other clinical outcomes of recipients under a new chemoprevention approach. This design also includes crucial implementation study outcomes for understanding the context and determinants of program outcomes (Figure 4). This design is suggested for cases where data on the effectiveness of the clinical intervention is needed, as we discuss later. Additionally, multiple stakeholders and factors are likely to influence these outcomes, such as national and sub-national leadership support, EPI and community health workers, cost-effectiveness, availability of case management, and funding for the hybrid deployment (supplementary Table S3).

**Figure 4.**
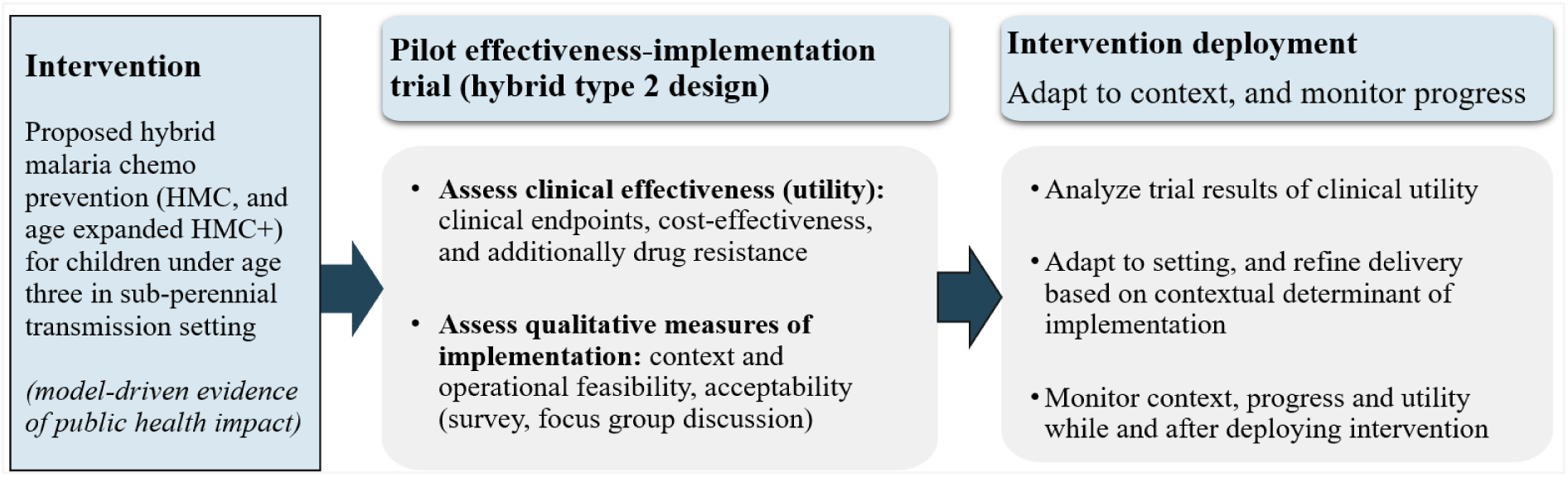
Potential implementation design for assessing the proposed hybrid chemoprevention dosing strategy in target setting.

## Discussion

Optimised chemoprevention strategies in sub-perennial malaria transmission settings have received little attention. We aimed to initiate multi-stakeholder dialogues to support chemoprevention guidelines for these settings by providing estimates of the potential public health impact and highlighting missed opportunities to protect vulnerable children. Our study used a validated open-access malaria model to [<u>15</u>, <u>19</u>] explore the potential public health benefits of complementing current PMC recommendations with additional seasonally-targeted monthly SP doses during the high-risk transmission period. Since, there is no practical experience deploying mixed malaria chemoprevention to children, this study aims to generate quantitative evidence for planning new dosing strategies. Furthermore, enhancing the public health impact of PMC will be beneficial in ensuring a wider uptake of this historically less utilized yet efficacious and safe intervention across recommended settings that also include sub-perennial seasonality[<u>1</u>]. We hope these early estimates of the potential impact, benefits or risks support conversations on targeted chemoprevention in these settings and support plans to generate empirical safety, feasibility, and impact data through pilot implementation studies.

We modelled a proposed hybrid malaria chemoprevention (HMC) strategy to cover children up to 24 months of age, which is aligned with the WHO’s PMC recommendations and consistent with experience to date[<u>1</u>]. However, since children remain vulnerable to severe malaria, and additional EPI contacts likely remain available up to 36 months of age, we also assessed an age-expanded HMC+ schedule. In addition, if these additional EPI contact points were utilised as malaria chemoprevention touchpoints, they may improve access to other vaccines or health services. We assumed different SP-sensitivities to account for resistance and ranges of access to case management and coverage levels to predict likely public health impact across example implementation scenarios and archetypal transmission settings. The HMC+ included age-expanded PMC+, which demonstrated a likely increase in net public health benefits and cost-effectiveness than current PMC[<u>25</u>] The monthly SP doses were given during the rainy season, combined with PMC or PMC+ throughout the year, to prevent interruption of current practices. Given the WHO guidelines on PMC dosing frequency, we considered a maximum of seven or nine PMC or PMC+ doses spread over two to three years[<u>1</u>]. However, children are likely to miss some doses during the rainy season when seasonally-targeted doses are administered to ensure a one-month gap between any two SP doses. These schedules were modelled over a wide range of prevalence settings (*Pf*PR_2-10_ 5-70%) in a combination of different drug sensitivity, healthcare strength (i.e. varying access to case management), and coverage assumptions. The model estimated efficacy against clinical, and severe cases, and the likely mode of parasite life stage activity of SP was validated to empirical data from randomized controlled trials and to a recent meta-data[<u>20</u>, <u>27</u>] as described previously[<u>25</u>].

We predicted that HMC and HMC+ would substantially increase protection by averting clinical and severe malaria burden compared to the current PMC alone. This is due to the greater chances of getting an SP dose, through additional seasonal dosing, during a higher-risk period, regardless of coverage. In contrast, unless a child’s age aligns with the timing of high-risk periods, PMC does not ensure protection. We also confirmed increased protection and maintenance of effectiveness in partially SP-resistant settings, albeit at a slightly reduced total malaria averted, also in line with earlier findings[<u>19</u>, <u>28</u>, <u>30</u>]. Although systematic reviews have confirmed that the effect of SP resistance is modest on the effectiveness of chemoprevention[<u>30</u>]. This implies that it would be prudent to monitor the evolution and spread of drug resistance by genetic biomarker surveys following continued and new chemoprevention. A recent case-control study found that a higher malaria burden may also be attributed to suboptimal drug concentration, possibly caused by missed doses rather than drug resistance[<u>31</u>]. Thus, alternative drug candidates must be carefully investigated to safeguard their chemoprevention benefits without compromising treatment options.

We also analyzed malaria outcomes from our simulations for children older than those covered by the proposed HMC or HMC+ (up to five years of age) to assess any post-intervention effects. Our results demonstrated a substantially larger positive net impact (cumulative cases by age during and in the post-intervention period) of both HMC and HMC+ compared to PMC alone, thus reducing potential delayed malaria burden. This positive net impact of HMC and HMC+ remained higher than PMC alone, even for scenarios with lower coverage. Notably, improved and reliable access to treatment will be necessary to ameliorate any increased risk of malaria and manage severe malaria cases after children are no longer protected by chemoprevention as is the case for any child who is no long using an effective form of malaria prevention). This is aligned with the WHO’s emphasis on strengthening healthcare systems. The increased impact of these chemoprevention strategies not only eases the demand on malaria treatment but also strengthens the capacity to prevent and treat other health needs[<u>32</u>]. Overall, these results alleviate concerns about delayed malaria.

To avoid potential safety concerns related to multiple dosing, we proposed HMC or HMC+ schedules that ensured a restriction for the number of doses each child received to ensure a one-month gap between any consecutive age- or seasonally-targeted SP doses. However, it will be necessary to integrate stakeholder, community, and implementation perspectives in planning the timing of hybrid dosing strategies, including the total number and timing of both age- and seasonally-targeted doses to understand the feasibility, acceptability, safety, and effective implementation and communication[<u>1</u>].

Additionally, we acknowledge the uncertainties related to changing climate conditions. For example, less rainfall in some years may lead to less seasonal variation in malaria transmission. Thus, we examined the potential benefit of rolling out HMC or HMC+ in both representative sub-perennial and perennial settings. As anticipated, the added benefit was larger in sub-perennial settings. However, the favourable net impact of the proposed hybrid schedule was also predicted in perennial settings. These results confirm that regardless of year-on-year rainfall and transmission uncertainty, hybrid chemoprevention delivery will likely increase the effectiveness.

Finally, we acknowledge that conducting implementation studies in real-world scenarios is crucial to translating our results into policy or recommendations. As discussed, results from seasonally-targeted IPTi trial in Senegal[<u>6</u>] and modelling results built on IPTi trial data from Ghana[<u>8</u>] indicated a substantially larger impact of seasonally-targeted SP over a decade ago. However, no follow-up investigation or implementation occurred. Data collection for formal implementation research is beyond the scope of our study; nevertheless, we aimed to address some likely next steps. We hope this facilitates broader discussion to ensure that new chemoprevention strategies advance beyond theoretical analysis and contribute to reducing the malaria burden in practice. To do this, we presented possible implementation designs[<u>13</u>] and potential determinants of implementation for the proposed HMC[<u>14</u>] based on the literature.

Although SP is a standard of care when given as PMC, data still needs to be generated to understand the effectiveness, feasibility, and the costs of alternative delivery strategies to support resource-allocation decisions. Therefore, hybrid type 2 effectiveness-implementation research will likely be helpful. We argue against the need for safety and efficacy studies given what’s known about safety & efficacy of individual doses of SP, that each dose protects for a certain period of time (depending on the resistance context) and that severe adverse reactions (primarily Stevens-Johnson syndrome) are idiosyncratic rather than dose-dependent[<u>33</u>]. Hence, evaluating effectiveness, consolidating safety and understanding impact is important. Furthermore, collecting qualitative data for the contextual determinants for tailoring delivery strategies to local contexts will be crucial to understanding how to begin rolling out any new delivery schedule. However, alternative study designs may also be considered, such as hybrid type 1 effectiveness-implementation with a primary emphasis on assessing clinical effectiveness and modest refinement to record the secondary implementation research goals. Program success will depend on the availability of funds to deploy additional staffing at EPI facility and on the ability of community health workers to deliver both age- and seasonally-targeted dosing.

As with all modelling studies, our results have several limitations. First, our predictions are based on a model of blood-stage parasite-clearing activity for SP[<u>25</u>] However, pyrimethamine may include liver-stage action, and we have yet to explore differences in immunity acquisition if any alternative mode of drug action dominates[<u>25</u>, <u>34</u>]. Also, our model does not estimate additional secondary benefits of SP beyond antimalarial effect (such as on bacterial or fungal infections)[<u>35</u>], which will further increase the total effect of HMC and HMC+. Second, our model does not estimate output by gender, and limited data on chemoprevention disaggregated by gender exists. Nevertheless, our PK/PD assumptions are consistent with earlier studies[<u>25</u>, <u>34</u>] and we expect the broad conclusions regarding the impact of HMC and HMC+ will hold. Third, we modelled SP as used in PMC, though implementation might prefer SP-AQ during high-risk periods. However, mixing PMC with SMC would require careful consideration of different drug schedules[<u>1</u>]. Thus, we assessed proposed hybrid delivery schedules with SP only as a first step. SP is a relatively inexpensive, single-dose drug available within EPI and is likely to have better adherence compared to a three-day schedule Increasing SP use may increase pressure on resistance development and spread. Thus, genomic surveillance would be prudent. Fourth, since there is currently no quantitative definition of sub-perennial transmission setting[<u>4</u>], we followed a conservative threshold below strictly seasonal (i.e. less than 60% in consecutive four months period[<u>2</u>, <u>3</u>] to define a representative sub-perennial setting. However, the transmission pattern can be flatter depending on the context, reinforcing the need for a more precise definition. Finally, we aimed to explore the potential impact of HMC, so we explored one implementation strategy only to initiate the discussion around possible designs.

## Conclusions

Our mathematical and pharmacological modelling results demonstrated increased public health impact in children under three years of age, using a hybrid malaria chemoprevention approach across a wide range of sub-perennial and perennial malaria transmission settings. The increase in burden averted was driven mainly by the timing of seasonally-targeted SP and the total number of doses (such as by age-expansion) rather than other setting parameters or intervention characteristics. The additional public health benefits of the proposed HMC and HMC+ compared to PMC alone were also realised under partial SP-resistance and reduced coverage assumptions. Both HMC and HMC+ demonstrated a positive net impact until age five, indicating limited delayed or age-shifted malaria burden. Moreover, higher access to case management mitigated delayed malaria effects and increased overall net benefit against severe malaria.

The proposed introduction of seasonally-targeted doses in hybrid chemoprevention programs is unlikely to interfere with the implementation of existing PMC and, instead, is likely to increase their public health impact. However, implementation studies are needed to build on our model-driven insights and generate empirical data on clinical impact, implementation feasibility, costs and cost-effectiveness. Together, these results will pave the way for conversations to rethink malaria chemoprevention, informing deployment decisions[<u>1</u>, <u>36</u>] and unlocking its full potential to protect children at high risk of severe malaria in sub-perennial transmission settings.

## List of abbreviations

EC50: half-maximal effective concentration
EIR: entomological inoculation rate
EPI: expanded program of immunization
HMC: hybrid malaria chemoprevention
HMC+: age-expanded hybrid malaria chemoprevention
IPTi: intermittent preventive treatment in infants
PMC: perennial malaria chemoprevention
PMC+: age expanded perennial malaria chemoprevention
PK/PD: pharmacokinetic/pharmacodynamic
PPPM: per person per month
PPYAR: per person per year at risk
SMC: seasonal malaria chemoprevention
SP: sulfadoxine-pyrimethamine
*Pf*: *Plasmodium falciparum*
*Pf*PR_2-10_: *Plasmodium falciparum* prevalence in 2-10 year olds
WHO: world health organization.

## Declarations

### Ethics approval and consent to participate

Not applicable

### Consent for publication

Not applicable

### Availability of data and materials

The publicly available source code for the applied individual-based model (OpenMalaria) can be found at https://github.com/SwissTPH/openmalaria (https://doi.org/10.5281/zenodo.10534022), including a detailed documentation at https://github.com/SwissTPH/openmalaria/wiki. The archived version of the model simulation and data analysis codes are available at (doi: 10.5281/zenodo.13804293). The R scripts and source data used for the production of figures presented in this paper can be found at (doi: 10.5281/zenodo.13805282).

### Authors’ contributions

SS and MAP designed the study. SS developed and performed analyses, drafted the manuscript and prepared figures. DS and MAP validated the workflow, analyses, and results. All authors contributed to interpreting the results, and making edits to the draft and final manuscript and gave their approval for publication.

## Acknowledgements

We acknowledge support and advice from all members of the Disease Modelling Research unit of the Swiss Tropical and Public Health Institute and thank Daria Hofer for the project management support. We acknowledge advice from implementation science researchers at the Institute for Implementation Science in Health Care, University of Zurich, for supporting implementation study ideas as part of a certification coursework to SS. All analysis and calculations were performed at sciCORE (https://ood.scicore.unibas.ch/) scientific computing centre at the University of Basel.

## Supplementary materials

### 1. Methods

#### 1.1 Model of malaria transmission and control (OpenMalaria)

In this model-driven commentary, we explored potential added public health benefit of a proposed hybrid malaria chemoprevention (HMC) strategy by combining age- and seasonally-targeted sulphadoxine-pyrimethamine (SP) doses[<u>1</u>, <u>2</u>]. We applied OpenMalaria, an open-source, individual based model of malaria epidemiology and control[<u>3</u>, <u>4</u>].The full range of parameters was originally calibrated to historical epidemiological data as described previously[<u>3</u>, <u>5</u>], and recalibrated to the latest data[<u>6</u>]. The source code for the model is publicly available at https://github.com/SwissTPH/openmalaria,

Essentially, the transmission of *Plasmodium* parasites between a mosquito vector and human host leads to infection in the human. This is simulated as a discrete, stochastic process in which the asexual blood stage parasite density in human drives the time-course and characteristics of malaria pathophysiology. The parasite density can be reduced either by naturally acquired immunity (variant-specific or variant-transcending or adaptive in the absence of any interventions), or by antimalarial treatment. The degree and duration of an intervention’s effect depends on its modelled mechanism of action, and the type of infection sub-model used (Table S1). The incidence of clinical malaria (symptomatic, but uncomplicated), severe cases (defined as per World Health Organization - WHO definition), and malaria-attributable, or all-cause mortality are tracked in each five-day timestep over the baseline (pre-intervention), intervention and post-intervention periods[<u>7</u>, <u>8</u>].

The vector sub-model characterizes the parasite (*Plasmodium)* life cycle in mosquito vector and the probability of transmission to a human host in each (five-day) timesteps. The exposure in human is extrapolated from entomological inoculation rate (EIR) i.e. the number of infectious mosquito bites per year, and is seasonally forced by applying two-component Fourier transformation (https://swisstph.github.io/openmalaria/fourier). Here, we modelled a generic vectoral pattern of *Anopheles gambiae,* currently one of the most prevalent mosquito species across Africa, by simulating the entomological characteristics[<u>9</u>, <u>10</u>].

The (OpenMalaria) model ensemble comprises of different model variants to address a variety of public health research questions. These have been developed by adapting the original model to incorporate varying assumptions about malaria transmissions, disease biology, anti-malarial immunity acquisition and decay, pharmacological effects of interventions, and impact of comorbidity on disease dynamics, among others[<u>11</u>]. In this work, we applied Molineux’s within-host model variant which mechanistically describes the time course of infection within each individual human host (as such, progression from asymptomatic to clinical malaria and to either severe outcome(s) or recovery)[<u>12</u>]. This variant was used as it enables explicitly modelling population pharmacokinetic and pharmacodynamic (PK/PD) profile of chemoprevention or treatment drugs in the target population including varying assumption about drug sensitivity based on parasite genotypes. The brief description of key model components and their applications in this study are detailed in Table S1, while full model description can be found at https://github.com/SwissTPH/openmalaria/wiki.

**Table S1.** Overview of OpenMalaria model, adapted from previous publications[<u>2</u>, <u>13</u>, <u>14</u>].

| Key modelled processes | Description and underlying assumptions | References |
| --- | --- | --- |
| **Malaria disease biology and epidemiology** |  |  |
| Infection in individual human host | Exposure is extrapolated from number of infectious mosquito bites over the year, specified by the entomological inoculation rate (EIR)<br>Seasonality pattern characterized by the monthly distribution of EIR<br>Correlates to availability based on age-dependent body surface area | [3, 15] |
| Progression of infection determined by asexual parasite density and acquired anti-malarial immunity | Explicit mechanistic within-host model of asexual parasitaemia<br>Various natural immunity (innate, variant-specific, variant-transcending, adaptive can reduce asexual parasite density in human) acquired over multiple age-dependent infection history<br>Both pre-erythrocytic liver-stage and blood-stage immunity decays exponentially over time<br>Duration of infection follows a log-normal distribution obtained by fitting to malaria therapy dataset in the original model (collated by Marsh and Snow[16]) | [3, 15, 17-20] |
| Clinical cases, morbidity, mortality and anaemia | Clinical malaria episode depends on human host parasite density and their pyrogenic threshold, which evolves over time depending on the individual exposure history<br>Patent malaria detected by threshold based on the type of diagnostic used (for instance, 40 parasites/μL of blood by microscopy detection was specified in this study)<br>Infections can be uncomplicated clinical cases, or can evolve to severe cases, based on parasite density and the pyrogenic threshold in each individual host<br>Severe disease can also be induced by typical age-based comorbidities | [3, 4, 21, 22] |
|  | • A proportion of the severe cases leads to deaths, resulting in direct malaria-related mortality<br>• Indirect mortality classified as deaths in patients not diagnosed as malaria deaths but would not occur without a malaria exposure<br>• All-cause mortality captures deaths resulting from both direct, indirect malaria-related causes, as well as hospitalization deaths, and thereby captures also impact of comorbidity (such as anaemia) |  |
| **Transmission setting characteristics** |  |  |
| Population age structure | • Flexible and informed by health and demographic surveillance data from Tanzania | [4, 23] |
| Transmission seasonality | • Specified by monthly EIR distribution<br>• In the absence of interventions seasonality is reproduced each year | [15, 24] |
| Transmission from infected humans to mosquitoes | • The level of infectivity depends on the density of the sexual form of parasites (gametocyte densities) present in human hosts extrapolated from blood-stage parasite densities, including a lag period<br>• Mosquitoes become infectious follows a binomial distribution | [3, 25, 26] |
| Entomological setting | • Mosquito lifecycle and behaviour towards human and non-human hosts (such as biting and resting) is embedded in a dynamic entomological model capturing the cycle of mosquito oviposition<br>• Multiple vector species can be simulated simultaneously<br>• Each variant-specific parasite has its own multiplication rate drawn from a normal distribution with mean of 16 | [27] |
| **Intervention deployment characteristics** |  |  |
| Drugs and other therapeutic modalities | • Interventions (small molecule drugs, vaccines, monoclonal antibodies, etc.) can act at different parasite life cycle stages (either by impacting on survival and emergence of asexual liver-stage or killing blood-stage parasites, blocking transmission) | [20, 28] |
| Case management | Modelled through a decision tree-based approach, which determines treatment implications depending on the occurrence of clinical cases<br>Its representation includes specification of diagnostic tests, effects of treatment, case fatality, case sequelae, and cure rates, influenced by the diagnostic threshold and access to care | [29] |
| Intervention deployment characteristics (as applied to model chemoprevention,) | In addition to treatment of clinical malaria episodes, chemoprevention interventions were deployed in the model either:<br>Continuously: to individuals at pre-specified ages over a specified time, and with specified coverage level (such as, administering perennial malaria chemoprevention)<br>Timed: at specified time points (annually or over multiple years) to targeted groups over several cycles and at specified coverage levels, typically seasonally-targeted (such as, administering seasonally-targeted sulphadoxine-pyrimethamine)<br>Interventions can be deployed by enrolling individuals into cohorts and tracking cohort outcomes, facilitating clinical trial simulation | [2, 20, 30-32] |
| Vector control | Probability of infection in human host can be reduced by vector control interventions<br>Interventions include long-lasting insecticide-treated nets (ITN), indoor residual spraying (IRS), house screening, baited traps, mosquito repellents, and push-pull | [27] |
| **Simulation characteristics and model variants** |  |  |
| Timestep | Simulations are tracked for every 5-day timestep<br>Tracked output metrics (for instance, clinical cases) from each simulated scenario can be extracted per timestep (5 days), monthly, quarterly or yearly values | [4] |
| Model variants | Varying assumptions for immunity decay, treatment effect, and heterogeneity in transmission are captured in 14 model variants | [2, 19, 20] |
|  | In this study Molineux within-host model adapted including explicit PK/PD characteristic of the intervention drugs for chemoprevention drug sulphadoxine-pyrimethamine (SP), and treatment drug artemether-lumefantrine (AL) |  |

#### 1.2 Scenario design

All simulations were run using ten random seeds to capture stochasticity. Trends, and dispersions for all outcome matrices were reported by calculating the median, and interquartile range respectively, as shown to be more robust to outliers from non-normal stochastic distribution[<u>33</u>].

##### Seasonality characteristics

We modelled two archetypal transmission settings: a representative sub-perennial (as was recorded in the replicated trial site in Manhiça, Mozambique[<u>2</u>, <u>34</u>, <u>35</u>]), and constant perennial transmission. Original normalized values based on average rainfall across Mozambique resembled seasonal transmission[<u>1</u>, <u>35</u>]. Therefore, we adapted transmission intensity by adjusting the Fourier transformation coefficient (Table S2). The adjusted seasonality depicted an example of archetypal sub-perennial transmission where transmission in a consecutive five-month period was below 60%. This was done to demarcate from strictly seasonal transmission settings typically covered under seasonal malaria chemoprevention[<u>1</u>]. The perennial transmission was modelled by uniform distribution of entomological inoculation rate over the year.

**Table S2.** Overview of model parameters for adapting sub-perennial transmission.

| Seasonality | Entomological inoculation rate rotate angle | Coefficient a1 | Coefficient a2 | Coefficient b1 | Coefficient b2 |
| --- | --- | --- | --- | --- | --- |
| Mozambique (normalized rainfall) | 0 | 1.0414 | 0.1916 | 1.2188 | 0.1078 |
| Sub-perennial<br>(adjusted values<br>as modelled) |  | 0.6414 |  | 0.5188 |  |

#### 1.2 Determinants of implementation outcome

The Consolidated Framework of Implementation Research (CFIR) provides a guiding framework for systemic data collection from individuals who influence the implementation outcomes[<u>36</u>]. It essentially includes five major domains, as outlined in Table S3. Each domain comprises of several factors or constructs that determine the outcome of an implementation.

**Table S3.** Brief description of the CFIR domains and constructs, adapted to this project [<u>36</u>, <u>37</u>].

| Domain name | Domain definition | Construct names in each domain |
| --- | --- | --- |
| **INNOVATION**<br>(renamed in the<br>updated CFIR from<br>INTERVENTION)[36] | The “thing” or intervention that is being implemented (such as, a proposed malaria chemoprevention strategy) | 1. Innovation Source<br>2. Innovation Evidence-Base<br>3. Innovation Relative Advantage<br>4. Innovation Adaptability<br>5. Innovation Trialability<br>6. Innovation Complexity<br>7. Innovation Design<br>8. Innovation Cost |
| **OUTER SETTING** | The setting that hosts the inner setting (such as, an epidemiological or clinical setting).<br>Multiple outer settings or levels of outer setting may exists in a scenario. | 1. Policies & Laws<br>2. Local Conditions<br>3. Local Attitudes<br>4. Partnerships & Connections<br>5. Financing<br>6. Critical Incidents<br>7. External Pressure |
| **INNER SETTING** | The setting in which the innovation is actually implemented (such as, Expanded Program of Immunization (EPI) within a village level population) | 1. Structural Characteristics<br>2. Communications<br>3. Culture (e.g. Recipient-Centeredness, Deliverer-Centeredness)<br>4. Tension for Change<br>5. Available Resources (Funding, Space, Materials & Equipment)<br>6. Relative Priority<br>7. Incentive Systems<br>8. Access to Knowledge & Information |
| **INDIVIDUALS** | The roles and characteristics of individuals in the context of the implementation including the recipients of the innovation/intervention (such as, children receiving malaria chemoprevention, the EPI staff, national malaria control program officers) | 1. High-level Leaders<br>2. Mid-level Leaders<br>3. Opinion Leaders<br>4. Implementation Facilitators<br>5. Implementation Leads<br>6. Implementation Team Members<br>7. Other Implementation Support<br>8. Innovation Deliverers<br>9. Innovation Recipients |
| **IMPLEMENTATION PROCESS** | The strategies and list of activities that are applied to implement the innovation/intervention | 1. Teaming<br>2. Assessing Needs (of innovation deliverers and recipients)<br>3. Assessing Context<br>4. Planning |
|  |  | 5. Tailoring Strategies<br>6. Engaging (innovation deliverers and recipients)<br>7. Doing<br>8. Reflecting & Evaluating (on both the implementation and the innovation)<br>9. Adapting |

### 2. Supplemental results

**Figure S1.**
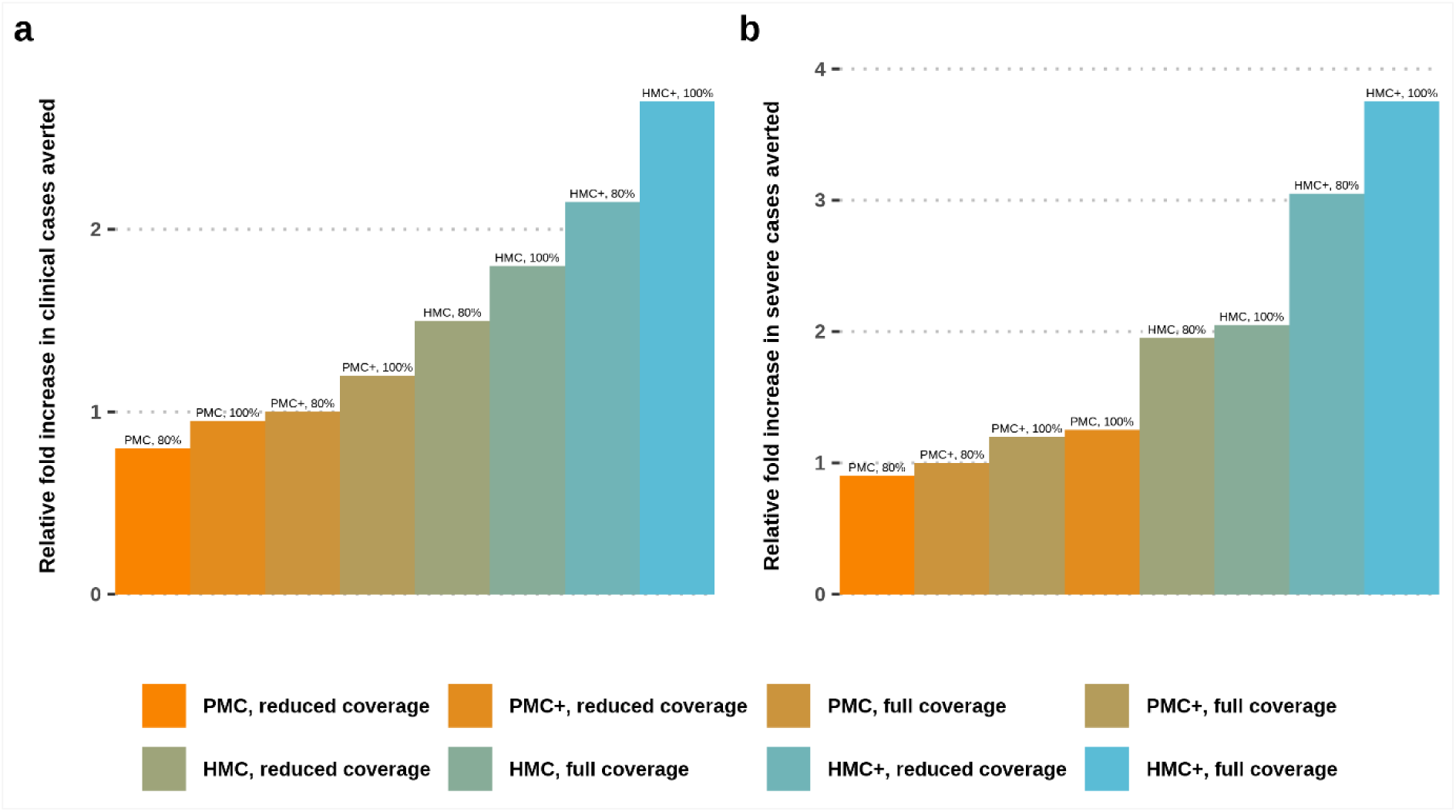
Median relative fold increase in burden averted by the proposed dosing schedules (HMC or HMC+) compared to PMC alone, against all episodes of clinical and severe malaria in first three years of life. The efficacy or effectiveness values were compared under full (100% in each dosing cycle) or reduced (80% in each dosing cycle) program coverage. The relative fold increase in burden averted by PMC+ compared to PMC is also depicted for a reference to the added benefit achieved by only age-expansion. Varying health system strength is represented by the low (10%), medium (30%) and high (50%) probability of accessing case management in a 14-day period post-diagnosis in medium to high transmission setting (*Pf*PR_2-10_ 30-39%, entomological inoculation rate 32). HMC: hybrid malaria chemoprevention. HMC+: age-expanded HMC; PMC: perennial malaria chemoprevention; PMC+: age-expanded perennial malaria chemoprevention; *Pf*PR2-10 : *Plasmodium falciparum* prevalence in children age 2-10.

**Figure S2.**
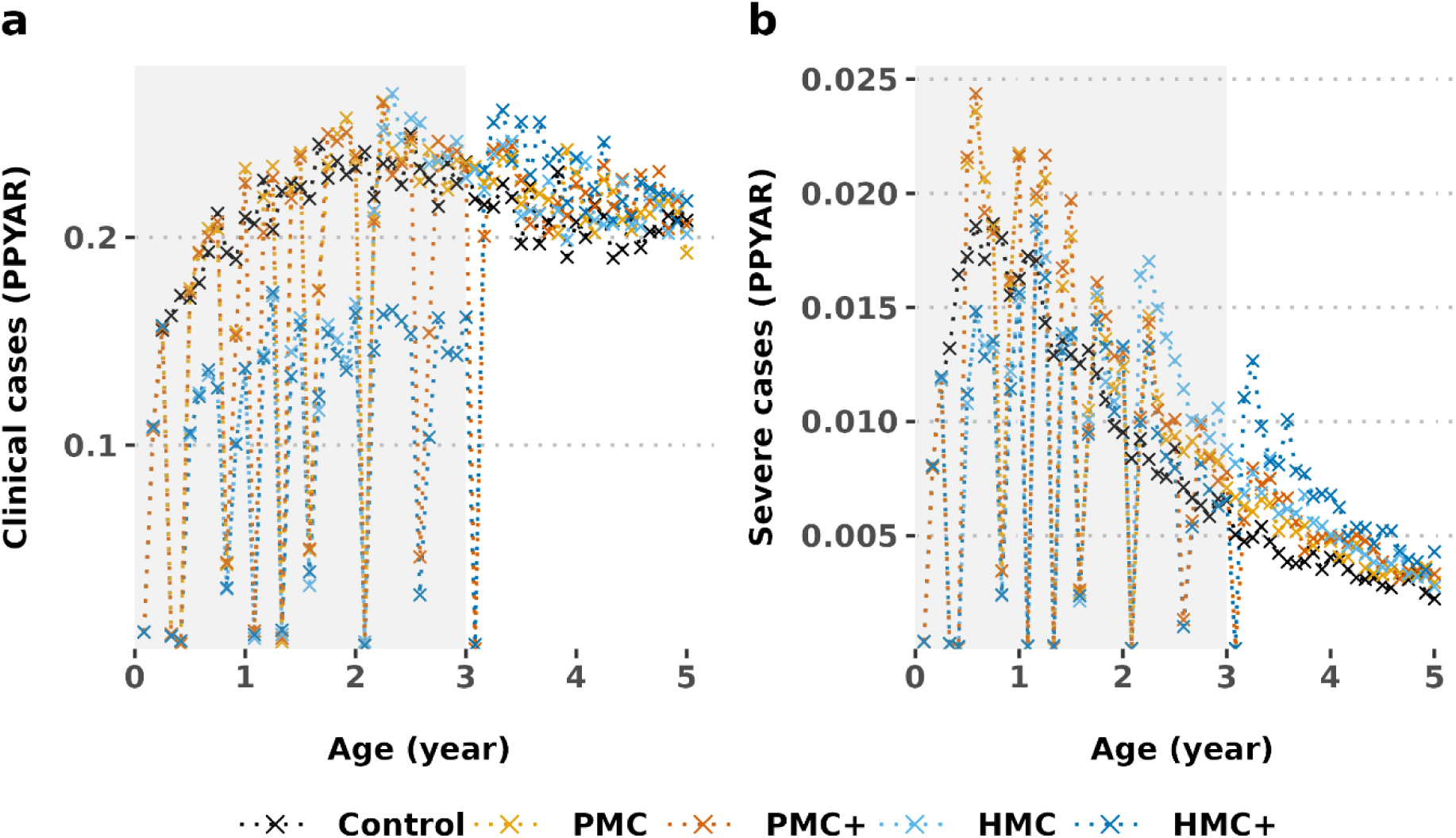
Age-pattern of clinical and severe malaria cases in intervention and follow-up ages. The grey shading denotes the intervention cohort. Results depicted in settings with *Pf*PR_2-10_ 30–39%, (entomological inoculation rate 32, 30% probability of access to care in a 14 day post-diagnosis) and 100% program coverage in partially SP-resistance (quadruple mutant in *Pfdhfr* and *Pfdhps* genes that reduces prophylactic protection to 35 days from 42 days in a sensitive setting). HMC: hybrid malaria chemoprevention. HMC+: age-expanded HMC; PMC: perennial malaria chemoprevention; PMC+: age-expanded perennial malaria chemoprevention; PPYAR: Per person per year at risk; SP: sulphadoxine-pyrimethamine.

**Table S3.**
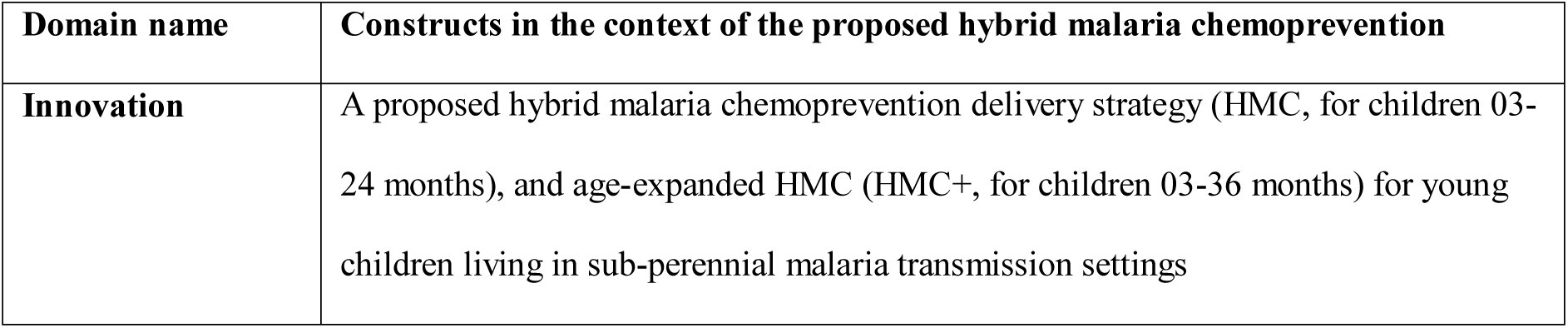
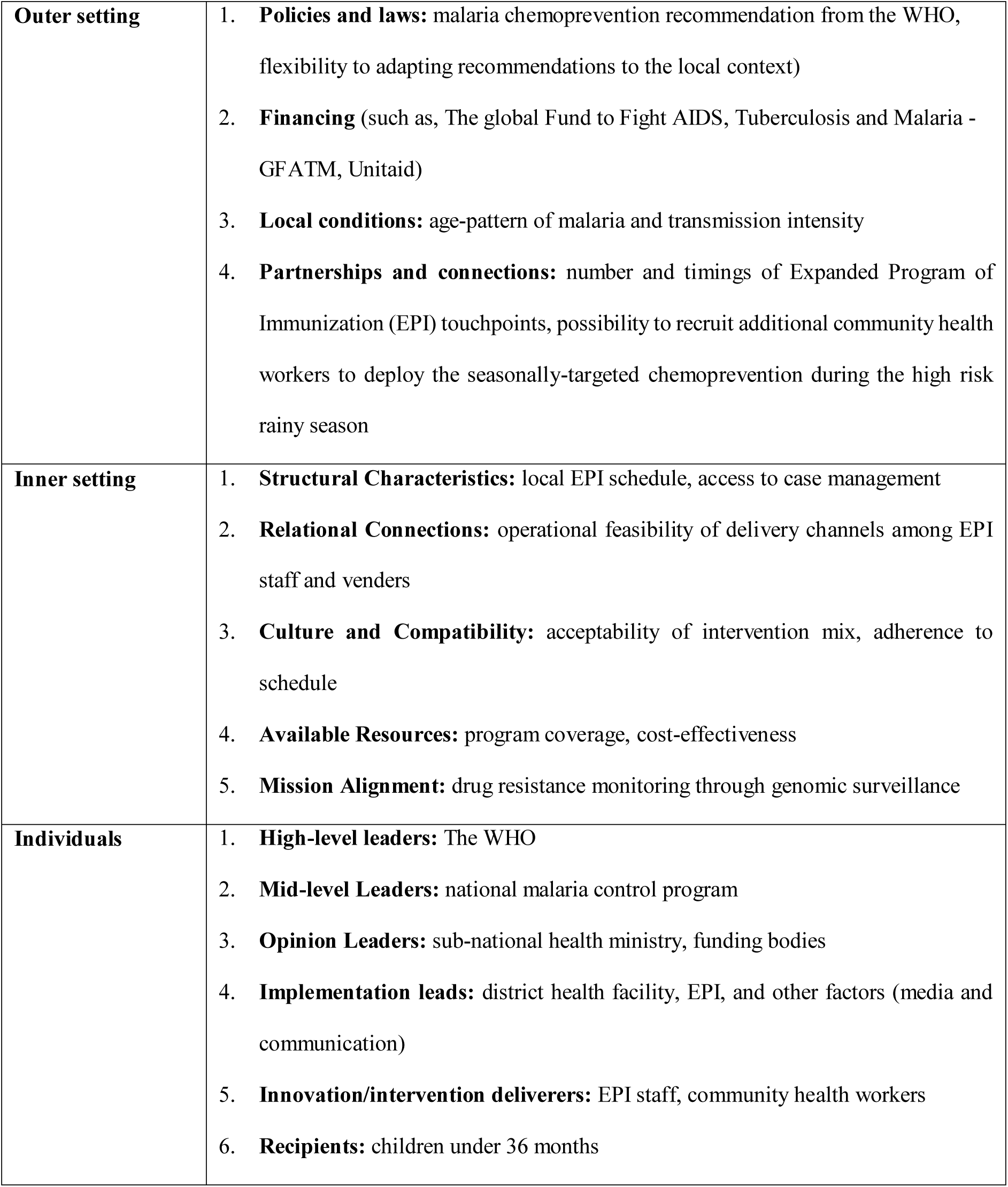
Potential determinants of implementation outcome as per Consolidated Framework for Implementation Research [<u>36</u>, <u>37</u>].

